# A cohort study of Post COVID-19 Condition across the Beta, Delta and Omicron waves in South Africa: 6-month follow up of hospitalised and non-hospitalised participants

**DOI:** 10.1101/2022.10.31.22281748

**Authors:** Waasila Jassat, Caroline Mudara, Caroline Vika, Richard Welch, Tracy Arendse, Murray Dryden, Lucille Blumberg, Natalie Mayet, Stefano Tempia, Arifa Parker, Jeremy Nel, Rubeshan Perumal, Michelle J. Groome, Francesca Conradie, Norbert Ndjeka, Louise Sigfrid, Laura Merson, Cheryl Cohen

## Abstract

**Background:** A third of people may experience persistent symptoms following COVID-19. With over 90% of South Africans having evidence of prior SARS-CoV-2 infection, it is likely that many people could be affected by Post COVID-19 Condition (PCC).

**Methods:** The was a prospective, longitudinal observational cohort study recruiting hospitalised and non-hospitalised participants, infected during the periods that Beta, Delta and Omicron BA.1 variants dominated in South Africa. Participants aged 18 years or older were randomly selected to undergo telephone assessment at 1, 3 and 6 months after hospital discharge or laboratory-confirmed SARS-CoV-2 infection. Participants were assessed using a standardised questionnaire for evaluation of symptoms and health-related quality of life. We used negative binomial regression models to determine factors associated with the presence of ≥1 symptoms at 6 months.

**Findings:** Among hospitalised and non-hospitalised participants, 46.7% (1,227/2,626) and 18.5% (199/1,074) had ≥1 symptoms at 6 months (p=<0.001). Among hospitalised participants 59.5%, 61.2% and 18.5% experienced ≥1 symptoms at 6 months among individuals infected during the Beta, Delta and Omicron dominant waves respectively. Among PLWH who were hospitalised, 40.4% had ≥1 symptoms at 6 months compared to 47.1% among HIV-uninfected participants (p=0.108).

Risk factors for PCC included older age, female sex, non-black race, the presence of a comorbidity, greater number of acute COVID-19 symptoms, hospitalisation/ COVID-19 severity and wave period (individuals infected during the Omicron-dominated wave had a lower risk of persistent symptoms [adjusted Incident Risk Ratio 0.45; 95% Confidence Interval 0.36 – 0.57] compared to those infected during the Beta-dominated wave). There were no associations between self-reported vaccination status before or after SARS-CoV-2 infection with persistent symptoms.

**Interpretation:** The study revealed a high prevalence of persistent symptoms among South African participants at 6 months although decreased risk for PCC among participants infected during the Omicron BA.1 wave. These findings have serious implications for countries with resource-constrained healthcare systems.

**Funding:** Bill & Melinda Gates Foundation, UK Foreign, Commonwealth & Development Office, and Wellcome.

## INTRODUCTION

Post COVID-19 Condition (PCC) as defined by the World Health Organization (WHO), “occurs in individuals with a history of probable or confirmed SARS-CoV-2 infection, usually 3 months from the onset of COVID-19 with symptoms that last for at least 2 months and cannot be explained by an alternative diagnosis” (1). Conservative estimates are that 10-30% people who have been infected with SARS-CoV-2 may be affected by PCC (2), while a study reported that as many as 60% of COVID-19 survivors will experience PCC at least during the first year (3). Furthermore, one study has reported persistent symptoms up to two years after acute COVID-19 (4). Systematic reviews have reported impaired functional status and reduced quality of life among adults with PCC (5-9).

PCC displays a wide spectrum of clinical manifestation and prevalence across different geographies (10). Over 60 physical and psychological sequelae have been described (6). The most common symptoms reported include fatigue, dyspnoea, arthromyalgia, headache, cough, chest pain, sleep disturbance, depression/anxiety, and cognitive deficits (‘brain fog’) including loss of memory and difficulty concentrating (3, 5, 11-13). Several studies identified higher rates of symptoms (4, 12) and higher rates of diabetes, respiratory and cardiovascular disease (12,14) in individuals with previous SARS-CoV-2 infection compared to general population and influenza controls.

Risk factors identified for PCC include female sex, ethnicity/race, specific comorbidities, greater number of acute COVID-19 symptoms, and severe COVID-19 disease requiring hospitalisation, ICU or invasive mechanical ventilation (5, 10, 15-18). Recent studies have suggested that the prevalence of PCC among people infected during the Omicron-dominant wave was lower than those infected in previous waves dominated by Delta and Alpha (10, 19-21). Some studies have also suggested that COVID-19 vaccination may reduce the risk of developing PCC (22-24). A single study suggested that the risk of PCC appears to increase with SARS-CoV-2 re-infection (25).

Existing studies on PCC are highly heterogeneous, with varying sample sizes, including patients with different acute COVID-19 severity and time frames for analysis (26). Many studies have not assessed risk factors, not examined the impact of SARS-CoV-2 variant, or have been conducted with a short follow-up duration. Most have been set in high-income settings (6, 27).

South Africa experienced five COVID-19 waves, dominated by the D614G mutation, Beta, Delta, Omicron BA.1 and Omicron BA.4/BA.5 variants, respectively. As of 6 September 2022, over 4 million cases (28), 542,332 hospitalisations and 104,302 deaths (29) have been reported. Nationally, 50% of adults have been fully vaccinated (30). Recent anti-SARS-CoV-2 antibody sero-prevalence surveys conducted just prior to the fifth wave revealed that over 90% of South Africans have immunity, the majority of whom had an antibody profile consistent with prior SARS-CoV-2 infection (31-33). It is therefore likely that a large number of people could be affected by PCC in South Africa, a country with a strained public health system pre-pandemic, which poses a challenge for the delivery of multidisciplinary care for people with PCC.

To characterise and identify risk factors for developing PCC in the population in South Africa, we established a longitudinal cohort study, following up hospitalised and non-hospitalised patients for 6 months post-laboratory confirmed SARS-CoV-2 infection. This study was led by the National Institute for Communicable Diseases (NICD), as part of a global study coordinated by the International Severe Acute Respiratory and emerging Infections Consortium (ISARIC). A recent publication detailed the prevalence of and risk factors for PCC among hospitalised participants in this cohort at 3 months (17). This is the first study in South Africa to characterise PCC up to 6 months post-SARS-CoV-2 infection, among hospitalised and non-hospitalised participants, and the impact of acute severity, HIV, SARS-CoV-2 variants (Beta, Delta, and Omicron) and COVID-19 vaccination on the risk of developing PCC.

## METHODS

### Study design

This was a prospective, longitudinal observational cohort study using an ISARIC open-access tool that was locally adapted to follow up participants with laboratory-confirmed COVID-19 in South Africa for 12 months (34).

### Study population

At the start of this study little was known about the prevalence of PCC to enable sample size calculation, so we selected a sample of 1,000 participants per cohort. The study population included individuals 18 years and older, with a positive reverse transcriptase polymerase chain reaction (RT-PCR) assay or rapid antigen test for SARS-CoV-2, who had a recorded contact number. Both non-hospitalised and hospitalised patients were recruited. The first cohort included hospitalised individuals infected during the period November 2020 to July 2021 when the Beta and Delta variants dominated, and assessments were conducted at 1, 3 and 6 months. We added a cohort from the period December 2021 to February 2022 when Omicron BA.1 dominated, but due to the need to obtain approvals, only 3- and 6-months assessments were conducted. A cohort of non-hospitalised individuals was recruited from the period August to November 2021 when the Delta variant dominated but again due to the need for approvals, only 3- and 6-month surveys were conducted for this cohort.

Non-hospitalised patients were identified through the national case line list (NMC-SS). Hospitalised patients were identified from public and private hospitals from all provinces of South Africa through the DATCOV national hospital surveillance. Of all eligible adults, a random sample were invited to participate in this study by telephone. Random sampling was done using a computer-generated list of eligible participants. If selected participants were not available, they were re-contacted twice until they were excluded. Verbal consent was obtained and recorded, and, where possible, assessments were conducted by trained researchers in the language of the participants’ choice.

### Measurements and instruments

A standardised case report form (CRF) and follow-up protocol developed by ISARIC, documented demographic variables, comorbidities, COVID-19 vaccination status, acute symptoms and severity, current health status, and new or persistent symptoms. The CRF contained validated tools to establish quality of life (EQ-5D-5L) (35), dyspnoea (modified MRC dyspnoea scale) (36) and functioning (UN/Washington Disability Score) (37). COVID-19 vaccination status was self-reported by participants and categorised as not vaccinated, or fully vaccinated before or after SARS-CoV-2 infection. (Individuals were considered to be fully vaccinated if they received two doses of BNT162b or one dose of Ad26.COV2.S with the most recent dose at least 14 days earlier and unvaccinated if these criteria were not met. These are the only two vaccines provided in the South African COVID-19 vaccination programme.)

The WHO COVID-19 clinical progression scale captured the range of clinical manifestations of patients with acute-COVID-19 (38). These scales were adapted to categorise levels of severity relevant to the in-hospital and non-hospitalised cohorts: (1) non-hospitalised asymptomatic; (2) non-hospitalised symptomatic; (3) hospitalised (no oxygen therapy); (4) hospitalised (oxygen by nasal prongs or mask); (5) hospitalised (mechanical ventilation and/or ICU).

Data was entered and stored on a secure online Research Electronic Data Capture repository (REDCap, version 10.6.14, Vanderbilt University, Nashville, Tenn.) hosted by the University of Oxford on behalf of ISARIC.

### Statistical analysis

Frequencies and percentages were used to summarise categorical data, and continuous data were expressed using medians and interquartile ranges (IQR). Frequency distribution tables and graphs were used to describe demographic characteristics, comorbidities, the prevalence of symptoms, and changes in quality of life. Bivariate analysis was conducted to compare the proportions of participants with ≥1 symptoms and no symptoms by wave period of infection, hospitalisation status, COVID-19 severity, and HIV status, employing Pearson chi squared test.

Negative binomial regression was implemented to assess factors associated with persistent symptoms at 6-months. The outcome variable was the presence of ≥1 symptoms. Variables assessed for association in the multivariable model were age, sex, ethnicity, presence of comorbid conditions (asthma, diabetes, hypertension, chronic cardiac disease, chronic kidney disease, malignancy, tuberculosis, HIV and obesity), number of symptoms during acute infection, acute COVID-19 severity, COVID-19 vaccination status, and wave period. These variables were included in the model a priori based on clinical plausibility. Variables with p<0.2 in the univariable analysis were included for multivariable analysis. Manual backward elimination was implemented, and the final model selection was guided by minimisation of the Akaike information criterion (AIC) or Bayesian information criterion (BIC). Statistical significance for the multivariable analysis was assessed at p<0.05. Statistical analyses were performed using Stata software version 16 (StataCorp Limited, College Station, Texas, USA).

The study was approved by the University of the Witwatersrand Human Research Ethics Committee (HREC M201150).

### Role of the funding source

The funders of the study had no role in study design, data collection, data analysis, data interpretation, or writing of the report.

## RESULTS

Of the 142,935 hospitalised and 273,429 non-hospitalised individuals eligible for inclusion, 13,868 (9.7%) hospitalised and 3,498 (1.3%) non-hospitalised were randomly selected for enrolment. Of those contacted, 3,334 (24.0%) hospitalised and 1,351 (38.6%) non-hospitalised were able to be reached, consented to participate, and were enrolled in the study. Of the 3,334 hospitalised participants recruited at 1 month, 2,841 (85.2%) and 2,626 (78.8%) completed 3- and 6-month assessment. Of the 1,351 non-hospitalised participants recruited at 3 months, 1,074 (79.5%) completed 6-month assessment (Supplementary Figure S1).

### Characteristics of participants

The median age of the hospitalised participants was 49 [IQR 37-60] years; 1,491 (56.8%) were females. The non-hospitalised participants’ median age was 37 [IQR 28-47] years; 641 (59.7%) were females. A pre-existing comorbidity was present in 65.1% and 32.8% of hospitalised and non-hospitalised participants respectively. The most common comorbidities were hypertension (32.1%), diabetes (20.0 %) and obesity (18.1%) among hospitalised participants; and hypertension (14.1%), diabetes (5.4%) and HIV (45, 4.2%) among non-hospitalised participants (Table 1).

**Table 1:**
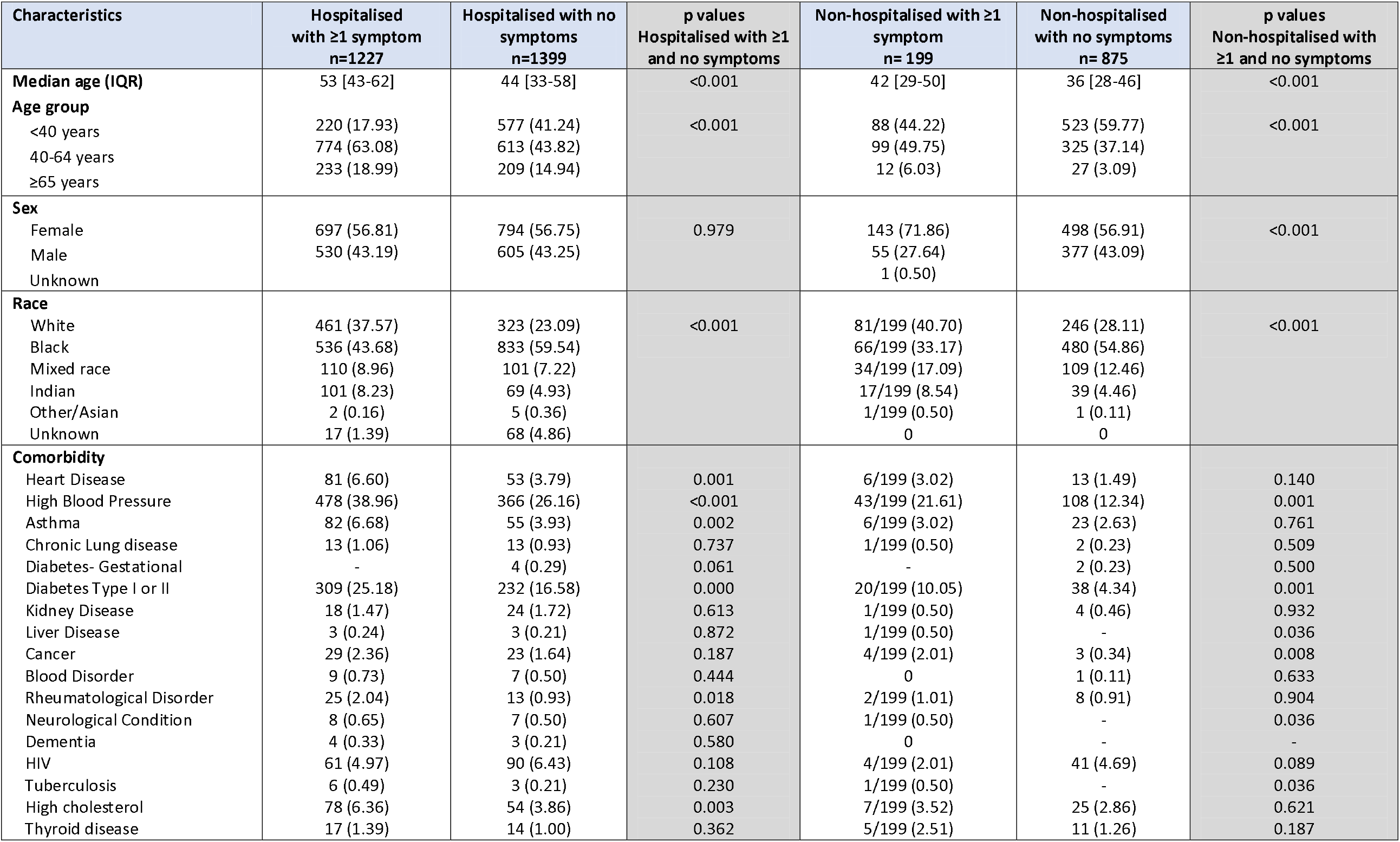
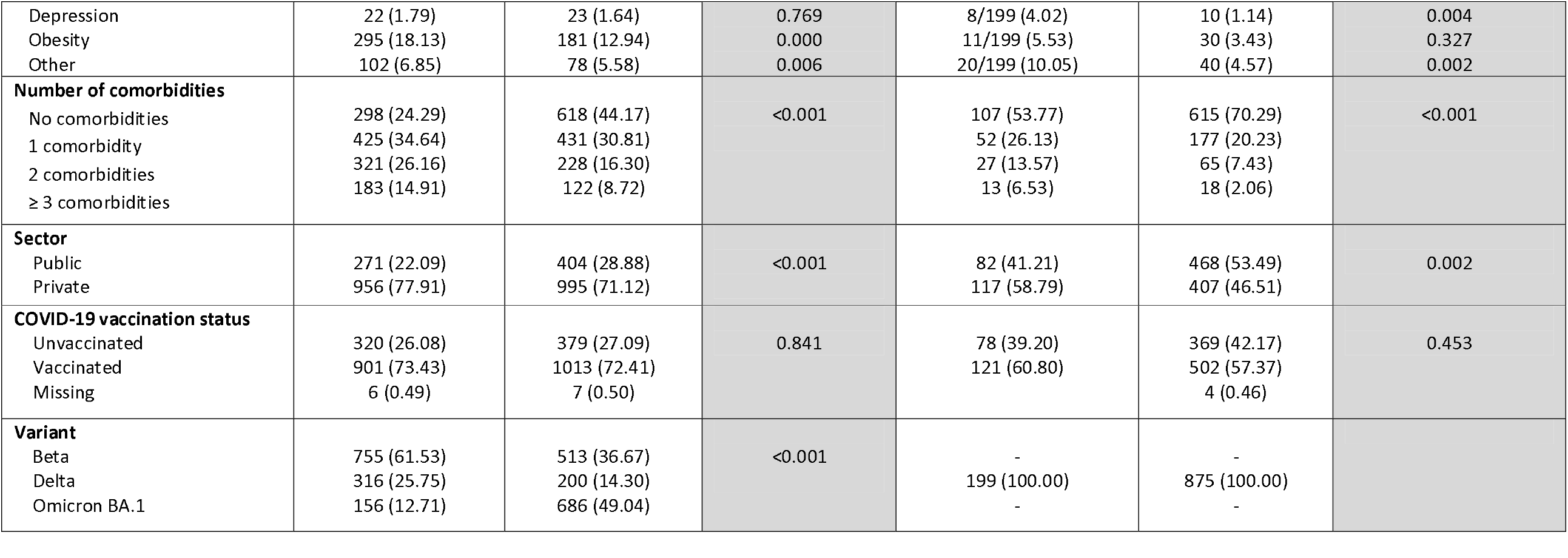
Characteristics of hospitalised and non-hospitalised participants with and without persistent symptoms, at 6-month follow-up

| Characteristics | Hospitalised with ≥1 symptom<br>n=1227 | Hospitalised with no symptoms<br>n=1399 | p values<br>Hospitalised with ≥1 and no symptoms | Non-hospitalised with ≥1 symptom<br>n= 199 | Non-hospitalised with no symptoms<br>n= 875 | p values<br>Non-hospitalised with ≥1 and no symptoms |
| --- | --- | --- | --- | --- | --- | --- |
| **Median age (IQR)** | 53 [43-62] | 44 [33-58] | <0.001 | 42 [29-50] | 36 [28-46] | <0.001 |
| **Age group** |  |  |  |  |  |  |
| <40 years | 220 (17.93) | 577 (41.24) | <0.001 | 88 (44.22) | 523 (59.77) | <0.001 |
| 40-64 years | 774 (63.08) | 613 (43.82) |  | 99 (49.75) | 325 (37.14) |  |
| ≥65 years | 233 (18.99) | 209 (14.94) |  | 12 (6.03) | 27 (3.09) |  |
| **Sex** |  |  |  |  |  |  |
| Female | 697 (56.81) | 794 (56.75) | 0.979 | 143 (71.86) | 498 (56.91) | <0.001 |
| Male | 530 (43.19) | 605 (43.25) |  | 55 (27.64) | 377 (43.09) |  |
| Unknown |  |  |  | 1 (0.50) |  |  |
| **Race** |  |  |  |  |  |  |
| White | 461 (37.57) | 323 (23.09) | <0.001 | 81/199 (40.70) | 246 (28.11) | <0.001 |
| Black | 536 (43.68) | 833 (59.54) |  | 66/199 (33.17) | 480 (54.86) |  |
| Mixed race | 110 (8.96) | 101 (7.22) |  | 34/199 (17.09) | 109 (12.46) |  |
| Indian | 101 (8.23) | 69 (4.93) |  | 17/199 (8.54) | 39 (4.46) |  |
| Other/Asian | 2 (0.16) | 5 (0.36) |  | 1/199 (0.50) | 1 (0.11) |  |
| Unknown | 17 (1.39) | 68 (4.86) |  | 0 | 0 |  |
| **Comorbidity** |  |  |  |  |  |  |
| Heart Disease | 81 (6.60) | 53 (3.79) | 0.001 | 6/199 (3.02) | 13 (1.49) | 0.140 |
| High Blood Pressure | 478 (38.96) | 366 (26.16) | <0.001 | 43/199 (21.61) | 108 (12.34) | 0.001 |
| Asthma | 82 (6.68) | 55 (3.93) | 0.002 | 6/199 (3.02) | 23 (2.63) | 0.761 |
| Chronic Lung disease | 13 (1.06) | 13 (0.93) | 0.737 | 1/199 (0.50) | 2 (0.23) | 0.509 |
| Diabetes- Gestational | - | 4 (0.29) | 0.061 | - | 2 (0.23) | 0.500 |
| Diabetes Type I or II | 309 (25.18) | 232 (16.58) | 0.000 | 20/199 (10.05) | 38 (4.34) | 0.001 |
| Kidney Disease | 18 (1.47) | 24 (1.72) | 0.613 | 1/199 (0.50) | 4 (0.46) | 0.932 |
| Liver Disease | 3 (0.24) | 3 (0.21) | 0.872 | 1/199 (0.50) | - | 0.036 |
| Cancer | 29 (2.36) | 23 (1.64) | 0.187 | 4/199 (2.01) | 3 (0.34) | 0.008 |
| Blood Disorder | 9 (0.73) | 7 (0.50) | 0.444 | 0 | 1 (0.11) | 0.633 |
| Rheumatological Disorder | 25 (2.04) | 13 (0.93) | 0.018 | 2/199 (1.01) | 8 (0.91) | 0.904 |
| Neurological Condition | 8 (0.65) | 7 (0.50) | 0.607 | 1/199 (0.50) | - | 0.036 |
| Dementia | 4 (0.33) | 3 (0.21) | 0.580 | 0 | - | - |
| HIV | 61 (4.97) | 90 (6.43) | 0.108 | 4/199 (2.01) | 41 (4.69) | 0.089 |
| Tuberculosis | 6 (0.49) | 3 (0.21) | 0.230 | 1/199 (0.50) | - | 0.036 |
| High cholesterol | 78 (6.36) | 54 (3.86) | 0.003 | 7/199 (3.52) | 25 (2.86) | 0.621 |
| Thyroid disease | 17 (1.39) | 14 (1.00) | 0.362 | 5/199 (2.51) | 11 (1.26) | 0.187 |
| Depression | 22 (1.79) | 23 (1.64) | 0.769 | 8/199 (4.02) | 10 (1.14) | 0.004 |
| Obesity | 295 (18.13) | 181 (12.94) | 0.000 | 11/199 (5.53) | 30 (3.43) | 0.327 |
| Other | 102 (6.85) | 78 (5.58) | 0.006 | 20/199 (10.05) | 40 (4.57) | 0.002 |
| **Number of comorbidities** |  |  |  |  |  |  |
| No comorbidities | 298 (24.29) | 618 (44.17) | <0.001 | 107 (53.77) | 615 (70.29) | <0.001 |
| 1 comorbidity | 425 (34.64) | 431 (30.81) |  | 52 (26.13) | 177 (20.23) |  |
| 2 comorbidities | 321 (26.16) | 228 (16.30) |  | 27 (13.57) | 65 (7.43) |  |
| ≥ 3 comorbidities | 183 (14.91) | 122 (8.72) |  | 13 (6.53) | 18 (2.06) |  |
| **Sector** |  |  |  |  |  |  |
| Public | 271 (22.09) | 404 (28.88) | <0.001 | 82 (41.21) | 468 (53.49) | 0.002 |
| Private | 956 (77.91) | 995 (71.12) |  | 117 (58.79) | 407 (46.51) |  |
| **COVID-19 vaccination status** |  |  |  |  |  |  |
| Unvaccinated | 320 (26.08) | 379 (27.09) | 0.841 | 78 (39.20) | 369 (42.17) | 0.453 |
| Vaccinated | 901 (73.43) | 1013 (72.41) |  | 121 (60.80) | 502 (57.37) |  |
| Missing | 6 (0.49) | 7 (0.50) |  |  | 4 (0.46) |  |
| **Variant** |  |  |  |  |  |  |
| Beta | 755 (61.53) | 513 (36.67) | <0.001 | - | - |  |
| Delta | 316 (25.75) | 200 (14.30) |  | 199 (100.00) | 875 (100.00) |  |
| Omicron BA.1 | 156 (12.71) | 686 (49.04) |  | - | - |  |

### Persistent symptoms in hospitalised versus non-hospitalised participants

Among hospitalised participants, 81.7% (1,932/2,366) had ≥1 symptoms at 1 month, 53.5% (1,518/2,840) at 3 months, and 46.7% (1,227/2,626) at 6 months (p=<0.001) (Figure 1). Among non-hospitalised participants, 31.6% (427/1,351) had ≥1 symptoms at 3 months, and 18.5% (199/1,074) at 6 months (p<0.001).

**Figure 1:**
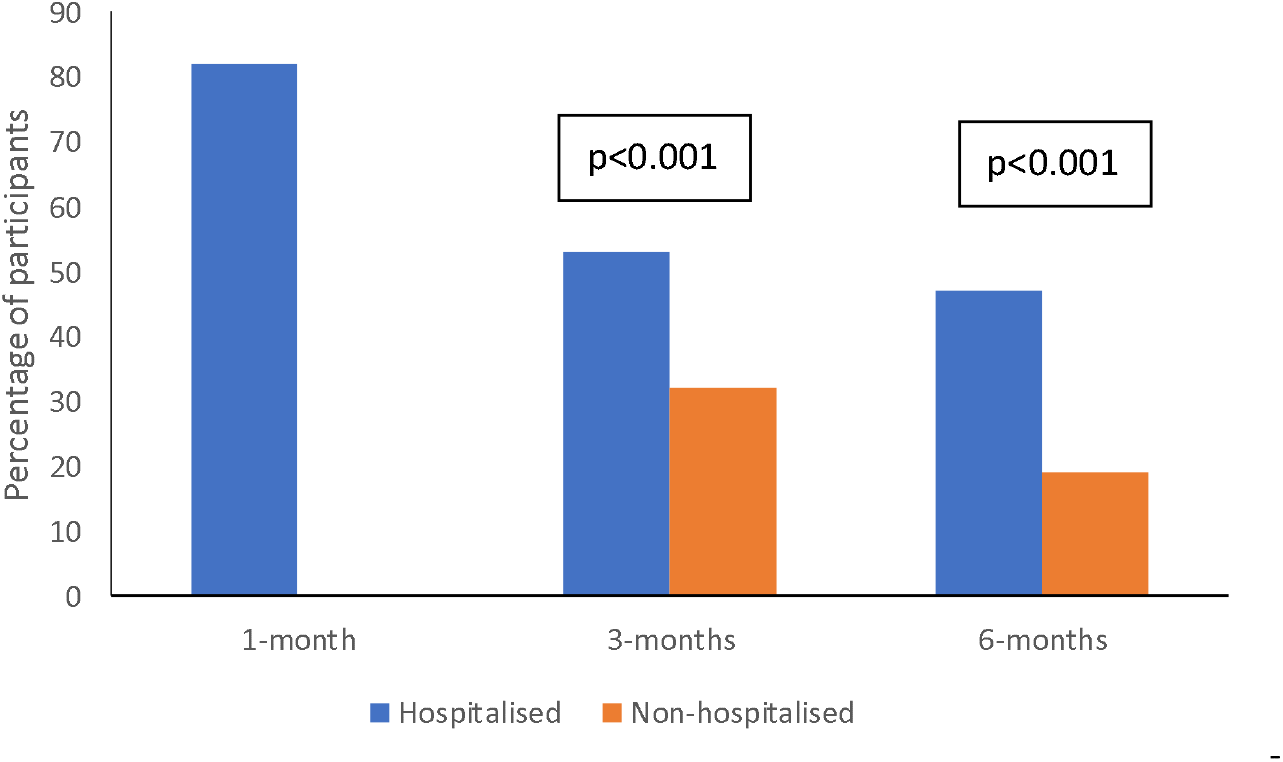
Percentage of hospitalised and non-hospitalised participants with ≥1 persistent symptoms, at 1, 3 and 6-months post-SARS-CoV-2 infection * assessments were not conducted at 1 month for non-hospitalised participants

The most common persistent symptoms reported among hospitalised participants at 6-months (n=2,626) were fatigue (32.1%), shortness of breath (15.6%), headache (10.3%), lack of concentration (9.9%) and muscle pain (8.5%) (Figure 2). All symptoms showed decreasing frequency from 1 to 6 months (Supplementary Figure S2). There were large decreases in the most common symptoms between 1 and 3 months, and little change, between 3 and 6 months.

**Figure 2:**
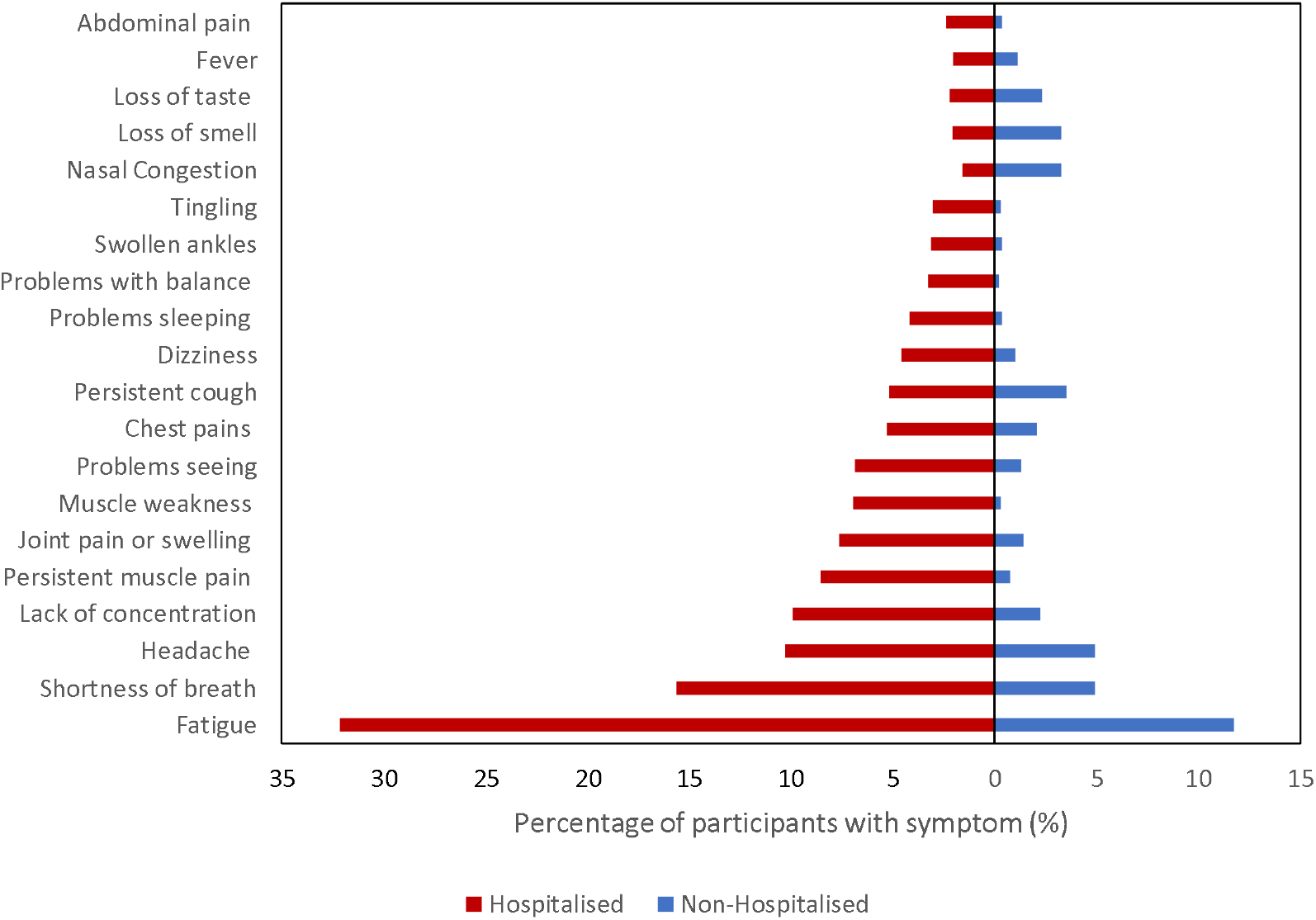
Frequency of most common symptoms among hospitalised and non-hospitalised participants at 6-months follow-up

The most common persistent symptoms reported among non-hospitalised participants at 6-months (n=1,074) were fatigue (11.7%), shortness of breath (4.9%), headache (4.9%), persistent cough (3.5%), loss of smell (3.3%) and nasal congestion (3.3%) (Figure 2). All symptoms showed decreasing frequency from 3 to 6 months (Supplementary Figure S2).

### Persistent symptoms by acute illness severity

Using an adapted WHO clinical scale of severity of the COVID-19 illness episode, persistent symptoms at 6-months were reported among 5.6% (9/162) of non-hospitalised asymptomatic participants, 20.8% (190/912) of non-hospitalised symptomatic participants, 17.1% (105/614) of hospitalised participants (no oxygen therapy), 51.1% (335/656) of hospitalised participants (received oxygen therapy), and 58.0% (787/1,356) hospitalised participants (received mechanical ventilation or treated in ICU) (p<0.001) (Supplementary Figure S3).

### Persistent symptoms among hospitalised participants by SARS-CoV-2 variant

The number of hospitalised participants included were 1,268, 516 and 842 admitted during the Beta, Delta and Omicron waves respectively. The median age of participants infected during the Beta wave was 52 years [IQR 41-61]; Delta wave 53 years [IQR 42-63] and Omicron wave 40 years [IQR 31-56] old. Most were females (Beta 51.7%, Delta 52.5%, Omicron 67.1%). A pre-existing comorbidity was present in 68.2%, 77.7% and 52.7% of participants admitted during the Beta, Delta and Omicron-waves respectively. (Supplementary Table S1).

Among participants infected during the Beta dominant wave, 77.5% (1,212) experienced ≥1 symptoms at 1 month, 64.3% (779) at 3 months, and 59.5% (755) at 6 months (p<0.001) (Figure 3). During the dominant Delta wave, 89.7% (720) ≥1 symptoms at 1 month, 71.2% (469) at 3 months, and 61.2% (316) at 6 months (p<0.001). During the Omicron dominant wave, 27.9% (270) had ≥1 symptoms at 3 months, and 18.5% (156) at 6 months (p<0.001).

**Figure 3:**
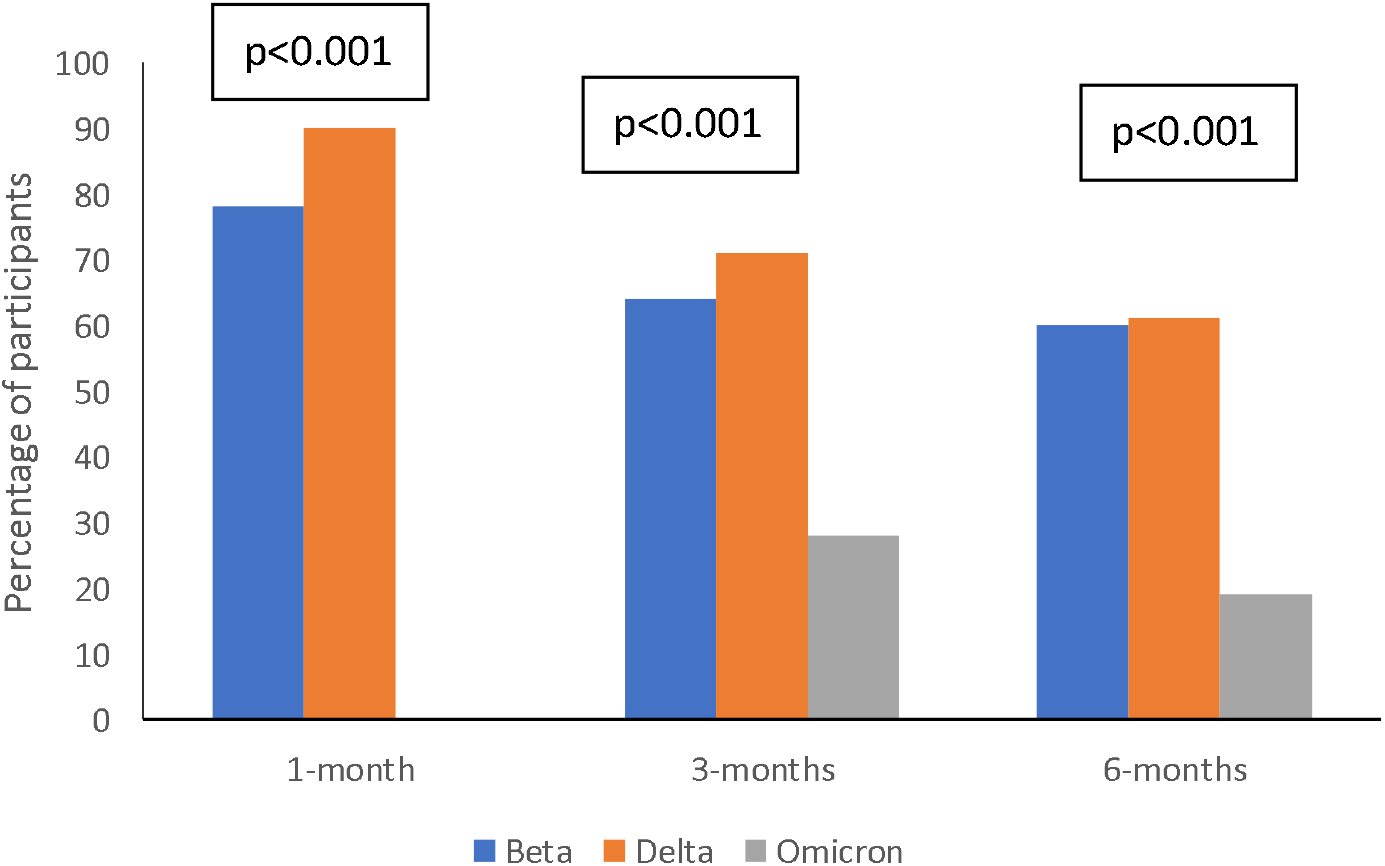
Percentage of hospitalised participants with ≥ 1 symptoms at 1, 3 and 6-month follow-up, by SARS-CoV-2 variant * assessments were not conducted at 1 month for participants infected during the Omicron wave

### Persistent symptoms amongst PLWH and HIV-uninfected participants

Among PLWH who were hospitalised, 43.1% (72/167) had ≥1 symptoms at 3 months, and 40.4% (61/151) at 6 months (p=0.624) (Supplementary Figure S4). Among HIV uninfected participants who were hospitalised, 54.1% (1,447/2,674) had symptoms at 3 months, and 47.1% (1,166/2,475) had symptoms at 6 months (p<0.001).

The most common persistent symptoms reported by PLWH (n=61) were fatigue (57.4%), shortness of breath (24.6%), headaches (21.3%), lack of concentration (18.0%) and muscle pain (14.8%).

### Symptom evolution amongst hospitalised participants

Amongst the 2,300 hospitalised participants who completed both the 3- and 6-months follow up assessment, the nature of symptom presentation and progression varied. While 32.6% (749) remained symptom-free throughout the follow-up period, 16.6% (381) experienced symptoms from the acute infection for 3 months, 31.2% (718) experienced symptoms from the acute infection until 6 months, 15.5% (357) experienced new symptoms at 3-months which continued to 6-months, and 4.7% (108) experienced new symptoms at the 6-months assessment (Supplementary Figure S5).

### Impact on quality of life amongst hospitalised participants

The proportion of hospitalised participants (N=2,626) who experienced reductions in quality of life domains at 6 months was 37.7% (989) for fatigue, 24.1% (633) for disability, 15.6% (410) for pain/discomfort, 13.4% (352) for anxiety/depression, 11.5% (303) for breathlessness, 8.3% (219) for usual activities, 7.6% (200) for mobility, and 2.6% (69) for self-care (Figure 4). Reported challenges with all the above quality of life measures decreased from 1, to 3, to 6 months for participants.

**Figure 4:**
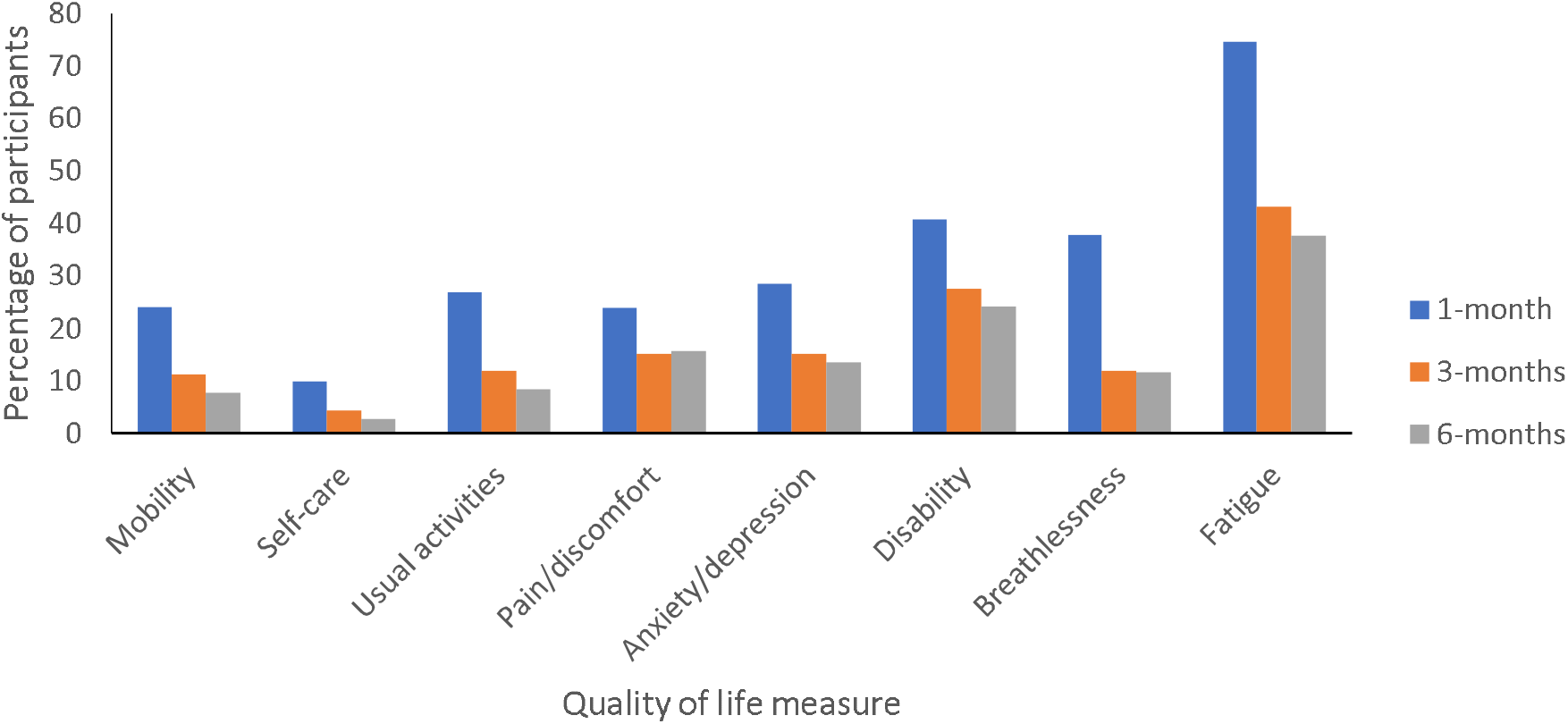
Percentage of hospitalised participants with decreased quality of life domains at 1, 3 and 6-month follow-up

### Factors associated with persistent symptoms

On multivariable regression, factors associated with ≥1 persistent symptoms among hospitalised and non-hospitalised participants at 6 months included age, sex, race, presence of a comorbidity, number of acute COVID-19 symptoms, COVID-19 severity, and SARS-CoV-2 variant (Table 2). There was an increased risk of persistent symptoms amongst older individuals aged 40-64 years (adjusted incident risk ratio [aIRR] 1.36, 95% confidence interval [CI] 1.18 – 1.57) and ≥65 years (aIRR 1.28; 95% CI 1.06 – 1.55) compared to those younger than 40 years; females (aIRR 1.24; 95% CI 1.12 – 1.38); and white (aIRR 1.26; 95% CI 1.11 – 1.38), Indian (aIRR 1.36; 95% CI 1.11 – 1.66), and mixed (aIRR 1.22; 95% CI 1.01 – 1.46) race compared to black race. Other risk factors included the presence of a comorbidity (aIRR 1.27; 95% CI 1.12 – 1.43); and 1-3 acute COVID-19 symptoms (aIRR 1.28; 95% CI 1.00 – 1.64) and 4 or more acute COVID-19 symptoms (aIRR 1.62; 95% CI 1.29 – 2.03) compared to no acute COVID-19 symptoms. With respect to severity of COVID-19, individuals who were non-hospitalised symptomatic (aIRR 2.32; 95% CI 1.15 – 4.70), hospitalised not on oxygen (aIRR 3.96; 95% CI 1.90 – 8.24), hospitalised on oxygen (aIRR 6.01; 95% CI 2.94 – 12.29), hospitalised ventilated or treated in ICU (aIRR 5.78; 95% CI 2.87 – 11.66) had a higher risk of persistent symptoms compared to non-hospitalised asymptomatic individuals. Individuals infected during the Omicron-dominated wave had a lower risk of persistent symptoms (aIRR 0.45; 95% CI 0.36 – 0.57) compared to those infected during the Beta-dominated wave. There were no associations between self-reported vaccination status before or after SARS-CoV-2 infection with persistent symptoms. Individual comorbidities including hypertension, diabetes, chronic pulmonary disease, obesity, HIV and others were not associated with persistent symptoms on negative binomial regression analysis.

**Table 2:** Multivariable regression of factors predicting ≥1 symptoms among hospitalised and non-hospitalised participants at 6 months after COVID-19.

| Characteristic | Persistent symptoms<br>n/N (%) | Unadjusted IRR (95% CI) | aIRR (95% CI) | p value |
| --- | --- | --- | --- | --- |
| **Age group (years)** |  |  |  |  |
| <40 | 308/1,408 (21.88) | Reference | Reference |  |
| 40-64 | 873/1,811 (48.21) | 2.20 (1.93 – 2.51) | 1.36 (1.18 – 1.57) | 0.0000 |
| $\geq 65$ | 245/481 (50.94) | 2.33 (1.96 – 2.75) | 1.28 (1.06 – 1.55) | 0.0095 |
| **Sex** |  |  |  |  |
| Male | 585/1,567 (37.33) | Reference | Reference |  |
| Female | 840/2,132 (39.40) | 1.06 (0.95 – 1.17) | 1.24 (1.12 – 1.38) | 0.0001 |
| Unknown | 1/1 (100) | 2.68 (0.38 – 19.05) | 3.33 (0.45 – 17.02) | 0.2317 |
| **Race** |  |  |  |  |
| White | 542/1,111 (48.78) | 1.55 (1.38 – 1.74) | 1.26 (1.11 – 1.38) | 0.0003 |
| Black | 602/1,915 (31.44) | Reference | Reference |  |
| Indian | 118/226 (52.21) | 1.66 (1.36 – 2.02) | 1.36 (1.11 – 1.66) | 0.0026 |
| Mixed race | 144/354 (40.68) | 1.29 (1.08 – 1.55) | 1.22 (1.01 – 1.46) | 0.0362 |
| Other | 3/9 (33.33) | 1.06 (0.34 – 3.30) | 1.23 (0.40 – 3.84) | 0.7198 |
| Unknown race | 17/85 (20.00) | 0.64 (0.39 – 1.03) | 1.08 (0.66 – 1.77) | 0.7653 |
| **Comorbidity** |  |  |  |  |
| No | 405/1,638 (24.73) | Reference | Reference |  |
| Yes | 1,021/2,062 (49.52) | 2.00 (1.78 – 2.25) | 1.27 (1.12 – 1.43) | 0.0002 |
| **Number of acute COVID-19 symptoms** |  |  |  |  |
| 0 | 103/671 (15.35) | Reference | Reference |  |
| 1-3 | 277/839 (33.02) | 2.15 (1.72 – 2.70) | 1.28 (1.00 – 1.64) | 0.0482 |
| $\geq 4$ | 1426/2190 (47.76) | 3.11 (2.54 – 3.81) | 1.62 (1.29 – 2.03) | 0.0000 |
| **COVID-19 severity** |  |  |  |  |
| Non-hospitalised asymptomatic | 9/162 (5.56) | Reference | Reference |  |
| Non-hospitalised symptomatic | 190/912 (20.83) | 3.75 (1.92 – 7.32) | 2.32 (1.15 – 4.70) | 0.0193 |
| Hosp (no oxygen) | 105/614 (17.10) | 3.08 (1.56 – 6.08) | 3.96 (1.90 – 8.24) | 0.0002 |
| Hosp (oxygen therapy) | 335/656 (51.07) | 9.19 (4.74 – 17.82) | 6.01 (2.94 – 12.29) | 0.0000 |
| Hosp (vent or ICU) | 787/1,356 (58.04) | 10.45 (5.42 – 20.15) | 5.78 (2.87 – 11.66) | 0.0000 |
| **Variant** |  |  |  |  |
| Beta | 755/1,268 (59.54) | Reference | Reference |  |
| Delta | 515/1,590 (32.39) | 0.54 (0.49 – 0.61) | 0.93 (0.81 – 1.07) | 0.3115 |
| Omicron | 156/842 (18.53) | 0.31 (0.26 – 0.37) | 0.45 (0.36 – 0.57) | 0.0000 |
| **COVID-19 vaccination status** |  |  |  |  |
| Not vaccinated | 398/1,146 (34.73) | 1.45 (1.21 – 1.73) | 0.97 (0.80 – 1.18) | 0.7544 |
| Vaccinated before infection | 175/731 (23.94) | Reference | Reference |  |
| Vaccinated after infection | 846/1,804 (46.90) | 1.96 (1.66 – 2.31) | 0.91 (0.75 – 1.10) | 0.3413 |
| Unknown | 7/19 (36.84) | 1.54 (0.72 – 3.28) | 0.81 (0.37 – 1.74) | 0.5815 |

\* aIRR: adjusted incident risk ratio; CI: confidence interval
The following individual comorbidities had p values <0.2 when assessed in univariate models: heart disease, asthma, cancer, rheumatological disorder, diabetes, hypertension and obesity. However, they were not significant in the multivariable model

## DISCUSSION

We described a high prevalence of PCC at 6 months following SARS-CoV-2 infection in a cohort of 3,700 hospitalised and non-hospitalised participants in South Africa. Overall, 39% of participants experienced persistent symptoms at 6 months; 46.7% in hospitalised participants and 18.5% in non-hospitalised participants. This differed by the variant period (lower prevalence of PCC among those infected during the Omicron period), and also the severity of acute COVID-19 (higher prevalence of PCC with more severe acute COVID-19).

The prevalence of PCC in hospitalised South African individuals (47%) was similar to estimates reported by other studies at 6 months, 40% in Italy (39), 40% in China (40), 47% in Switzerland (41), 48% in Saudi Arabia (42), 50% in Russia (43), 57% in US (44), 60% in France (45), 61% in Norway (46), 68% in China (47) and 64% in a meta-analysis (48). Differences in the prevalence of PCC by geographical region at 3-months have previously been reported, ranging from 31% in North America, to 44% in Europe, and 51% in Asia (10).

Encouragingly, the prevalence of persistent symptoms in our cohort declined with successive follow-up periods, from 82% at 1 month to 53% at 3 months and 47% at 6 months for hospitalised participants, and from 32% at 3 months to 19% at 6 months for non-hospitalised participants. A declining trend in persistent symptoms between 3 and 6 months has also been reported from 51% to 40% in a Chinese cohort (40), and from 68% to 60% in France (45). Other studies with longer follow-up durations have reporting further declining trends at 12 months (43,49) and 24 months (4). However, a significant proportion of participants in our study remained symptomatic at 6-months, representing a substantial potential individual and health system burden. Extrapolating the prevalence of PCC to the total hospitalised COVID-19 patients (from hospital surveillance) and likely total infected individuals (from serosurveys), it is possible that over 250,000 hospitalised and 6.3 million non-hospitalised individuals had persistent symptoms at 6 months after SARS-CoV-2 infection in South Africa.

We also showed that the progression of PCC was not linear and was characterised by fluctuating trends over the study period. This finding is consistent with the WHO’s description of PCC as a condition characterised by a clinical progression including new onset of symptoms following initial recovery from an acute COVID-19 episode, or persistent symptoms from the initial illness, or symptoms that fluctuate or relapse over time (1). Studies have described the relapsing and remitting nature of PCC (44, 50). However, few studies have explored the nature of symptom progression in the detail provided by this study.

Fatigue was the most common symptom in our study at all timepoints and amongst all cohorts. COVID-19 induced fatigue can be defined as ‘a decrease in physical and/or mental performance that results from changes in central, psychological, and or/peripheral factors due to the COVID-19 disease’ (51). Fatigue was reported to be the most debilitating PCC symptom, and the main reason patients contacted a COVID rehabilitation programme (52). Similar rates of fatigue, and decreased quality of life have been reported after the SARS and MERS epidemics (8).

Risk factors for PCC in our study included older age, female sex, non-black race, the presence of a comorbidity (although individual comorbidities were not predictive), greater number of acute COVID-19 symptoms, hospitalisation/ COVID-19 severity and wave period. Other studies have reported on associations of PCC with female sex, >5 symptoms during acute COVID-19, co-morbidities (pre-existing hypertension, chronic lung conditions, obesity, asthma), hospital admission, and severity of acute COVID-19 (5, 10, 15, 16, 18), as well as oxygen treatment (5, 53, 54), steroid treatment (54), and treatment in ICU (39). Conflicting findings have been reported with regards to age and race. Some studies reported increased risk with older age (5, 6, 11, 15, 18, 41, 55). Several studies have identified higher risk of PCC among adults less than 70 years old (34, 56), among young adults (16, 44, 57), and among middle-aged adults 40-55 years (3, 58). Some studies reported increased risk of PCC for ethnic minority groups (6, 55, 59), while other studies concluded that race and ethnicity were not associated with PCC (60).

The risk of persistent symptoms at 6 months was higher as severity of acute COVID-19 increased. Similar findings have been reported of higher burden of PCC amongst those hospitalised compared to non-hospitalised (61), and among those with severe acute COVID-19 in systematic reviews (5, 62) and cohort studies (63, 64). The mechanisms for increased risk of PCC with severity include more severe immune response and cytokine storm resulting in more organ damage, as well as more aggressive treatment (65).

We described a lower risk of persistent symptoms among participants infected in the Omicron wave. Similar findings have been reported in other studies with shorter follow-up duration (10, 19-21). It however remains unclear whether this reduction in PCC is related to the Omicron variant, or as a result of the effect of prior immunity from vaccination or natural infection, which resulted in milder acute COVID-19 infection.

A few studies have suggested that COVID-19 vaccination reduces the likelihood of PCC (22-24) and triggered improvement in symptoms (66, 67). The mechanisms suggested for this include destruction of residual viral reservoir by the antibody response, and a “reset” of autoimmune dysregulation (24, 53). In our study, participants who were vaccinated had no statistical difference in persistent symptoms compared to those who were unvaccinated. However, it is important to note that our study did not record objective vaccination status, details of partial versus complete vaccination, precise temporality to infection, or serological confirmation of immunity.

Finally, our study also demonstrated the significant impact of PCC on quality of life, however quality of life improved for all domains over time. The quality-of-life domains that were still significantly affected at 6 months were fatigue (38%), disability (24%), and pain/discomfort (16%). Other studies reported 37% (6), 29% (68) and 25% (69) of participants had a reduced quality of life overall. In a systematic review, all studies with follow-up of 3-6 months post COVID-19, demonstrated worsened mobility, self-care, usual activities, pain/discomfort, and anxiety/depression, resulting in loss of independence (9). Systematic reviews reported activity impairment, disability and disruption in work life (5, 7, 8, 13).

### Strengths and limitations

Our study was a large, nationally representative longitudinal cohort study and, to our knowledge, the first of its kind in South Africa and sub-Saharan Africa. As part of the global ISARIC collaboration, we used standardised and validated tools, which allows for comparison across participating countries.

The study had several limitations. First, we did not include controls with respiratory infection other than COVID-19 to understand the effect of COVID-19 on continuing morbidity. Secondly, all participants were enrolled through a telephone assessment, limiting the enrolment to those who had a telephone number recorded. Thirdly, there is a possibility of response bias, and participants who had symptoms might have been more likely to participate than those who did not. The high retention and minimal loss to follow-up in our study strengthens our findings (retention rate was 78.8% and 79.5% for hospitalised and non-hospitalised participants respectively at 6 months). Recall bias is possible as participants were recruited at 1 month or 3 months and asked to report their symptoms at acute COVID-19. Some bias could also have been introduced due to self-reporting of comorbidities and vaccination status. Finally, the lack of objective metrics to assess symptoms might affect reporting, which is a limitation of all PCC studies (41, 70), and future work should focus on developing objective assessments and examining their correlation with self-reported measures.

## CONCLUSION

The study revealed a high prevalence of PCC among South African participants at 6 months, but encouragingly showed a decreasing prevalence of persistent symptoms and impact on quality of life over time. The study improves our understanding of the nature and evolution of PCC, the risk factors associated with PCC in the South African context, and the differences in PCC by circulating SARS-CoV-2 VOCs. The finding of decreased PCC at 6 months among participants infected during the Omicron wave is important as it may suggest decreasing rates of PCC with greater population immunity.

These findings have serious implications for LMICs which have resource-constrained healthcare systems that may now need to also establish post-acute care services in settings where physical, cognitive, and mental health disabilities often go under-recognised (57). LMICs also do not generally have social safety nets, and the impact of chronic sequelae on the workforce and on families’ livelihoods remain a concern.

The evidence generated by this study will help to inform the national public health response to PCC, which should include sensitising and training health care workers on managing patients with PCC, updating clinical guidelines, and establishing multidisciplinary health services at least in large public hospitals. These findings will also inform the development of additional patient support forums and educational material for patients with PCC. The study continues to follow-up participants and will report on trends at the final survey to be conducted at 12 months.

## Data Availability

Restrictions apply to the availability of these data and so they are not publicly available. However, data can be made available from the corresponding author upon reasonable request and with the permission of the South African Department of Health and the NICD.

## Contributors

WJ had the idea for and designed the study, had full access to all the data in the study, and takes responsibility for the integrity of the data and the accuracy of the data analysis. WJ and CM drafted the paper. CM and RW did the analysis. All authors critically revised the manuscript for important intellectual content and agreed to submit the final version for publication. MD, CM, CV, TA and the research team completed the follow-up work. MD, CM, CV, TA and the research team collected and verified the data. All authors agree to be accountable for all aspects of the work in ensuring that questions related to the accuracy or integrity of any part of the work are appropriately investigated and resolved.

## Declaration of interests

We declare no competing interests.

## Acknowledgments

We would like to acknowledge all the members of the research team (namely Ashrina Kandier, Bawinile Hlela, Bibianna Chikowore, Claudette Kibasomba, Dorcas Magorimbo-Njanjeni, Jeniffer Nagudi, Kadija Shangase, Khutso Maphoto, Lilford Lesabe, Lindelwa Ngobeni, Manana Sibanda, Menzi Mbonambi, Molebogeng Moyo, Ncamsile Mavundla, Okaeng Plaatjie, Salaminah Mhlanga, Sinalo Gqunu, Tasmeeya Moola, Thandeka Kosana, and Zelna Jacobs) responsible for the recruitment of participants into the study and subsequent follow-up surveys. We are grateful to the Bill & Melinda Gates Foundation (especially Georgina Murphy and Keith Klugman) for their continued support of the project. We thank the ISARIC team (especially Daniel Plotkin) for help in developing the CRF and supporting the REDCap database. Moreover, we thank the members of the Long COVID support group who informed the CRF. Finally, we would like to acknowledge all members of NICD and the DATCOV Team who provided technical support and assisted with data management, and we express gratitude to all the patients who participated in the contribution to the understanding of PCC. This work was made possible by funding from the Bill & Melinda Gates Foundation (INV-008112 and INV-032725) and, through our collaboration with ISARIC, was supported by the UK Foreign, Commonwealth & Development Office, Wellcome (215091/Z/18/Z), and the Bill & Melinda Gates Foundation (OPP1209135).

## Notes

### Competing Interest Statement

The authors have declared no competing interest.

