## Supplementary Materials for "A cohort study of Post COVID-19 Condition across the Beta, Delta and Omicron waves in South Africa: 6-month follow up of hospitalised and non-hospitalised participants"

**SUPPLEMENTARY FIGURES AND TABLES**

**Enrolled hospitalised participants at 1 month N=3,334**

1,563 Beta wave

803 Delta wave

968 Omicron wave

17 died

48 unwilling to participate

428 unreachable

**Surveys completed at 6 months follow up N=2,626 (78.8%)**

1,268 Beta dominant wave

516 Delta dominant wave

842 Omicron dominant wave

17 died

42 unwilling to participate

155 unreachable

**Surveys completed at 3 months follow up N=2,841 (85.2%)**

1,214 Beta wave

659 Delta wave

968 Omicron wave

1 died

24 unwilling to participate

252 unreachable

**Enrolled non hospitalised participants at 3 months N=1,351**

1,351 Delta dominant wave

**Surveys completed at 6 months follow up N=1,074 (79.5%)**

1,074 Delta dominant wave

Supplementary Figure S1: Study population based on inclusion criteria

Percentage of participants

|  | 1 month | 3 months | 6 months |
| --- | --- | --- | --- |
| Hospitalised Fatigue | 66 | 39 | 32 |
| Hospitalised SOB | 48 | 19 | 16 |
| Hospitalised Headache | 22 | 14 | 10 |
| Hospitalised Muscle weakness | 21 | 19 | 9 |
| Hospitalised Lack of concentration | 18 | 12 | 10 |
| Non-hospitalised Headache |  | 7 | 5 |
| Non-hospitalised Fatigue |  | 19 | 12 |
| Non-hospitalised SOB |  | 3 | 5 |
| Non-hospitalised Cough |  | 3 | 4 |
| Non-hospitalised back pain |  | 5 | 1 |
| Non-hospitalised Loss of smell |  | 5 | 3 |
| Non-hospitalised Nasal congestion |  | 3 | 3 |

Supplementary Figure S2: Frequency of most common symptoms among hospitalised and non-hospitalised participants at 1, 3 and 6-months follow-up

Percentage of participants

|  | Persistent symptoms |
| --- | --- |
| Non-hospitalised asymptomatic | 6 |
| Non-hospitalised symptomatic | 21 |
| Hospitalised (no oxygen therapy) | 17 |
| Hospitalised (oxygen therapy) | 51 |
| Hospitalised (mechanical ventilation or ICU) | 58 |

Supplementary Figure S3: Percentage of participants with ≥1 symptoms at 6-month follow-up, by acute COVID-19 severity

Supplementary Table S1: Characteristics of hospitalised participants at 6-month follow-up, by variant

| **Category** | **Beta (n=1,268)** | **Delta (n=516)** | **Omicron (n=842)** | **p values** |
| --- | --- | --- | --- | --- |
| **Median age in years (IQR)**  **Age group**  <40 years  40-64 years  ≥65 years | 52 [41-61]  292 (23.03)  763 (60.17)  213 (16.80) | 53 [42-63]  98 (18.99)  306 (59.30)  112 (21.71) | 40 [31-56]  407 (48.34)  318 (37.77)  117 (13.90) | <0.001  <0.001 |
| **Sex**  Female  Male | 655 (51.66)  613 (48.34) | 271 (52.52)  245 (47.48) | 565 (67.10)  277 (32.900 | <0.001 |
| **Race**  White  Black  Mixed race  Indian  Other  Unknown | 363 (28.63)  686 (54.10)  115 (9.07)  93 (7.33)  1 (0.08)  10 (0.79) | 264 (51.16)  171 (33.14)  45 (8.72)  34 (6.59)  1 (0.19)  1 (0.19) | 154 (18.29)  512 (60.81)  49 (5.82)  43 (5.11)  5 (0.59)  79 (9.38) | <0.001 |
| **Comorbidity**  Heart Disease  High Blood Pressure  Asthma  Chronic Lung disease  Diabetes- Gestational  Diabetes Type I or II  Kidney Disease  Liver Disease  Cancer  Blood Disorder  Rheumatological Disorder  Neurological Condition  Dementia  HIV  Tuberculosis  High cholesterol  Thyroid disease  Depression  Obesity  Other | 60 (4.73)  451 (35.57)  61 (4.81)  12 (0.95)  0  311 (24.53)  16 (1.26)  2 (0.16)  13 (1.03)  7 (0.55)  16 (1.26)  7 (0.55)  3 (0.24)  74 (5.84)  3 (0.24)  60 (4.73)  13 (1.03)  10 (0.79)  269 (21.21)  52 (4.10) | 42 (8.14)  205 (39.73)  28 (5.43)  7 (1.36)  1 (0.19)  90 (17.44)  11 (2.13)  2 (0.39)  14 (2.71)  4 (0.78)  8 (1.55)  3 (0.58)  2 (0.39)  16 (3.10)  3 (0.58)  37 (7.17)  11 (2.13)  11 (2.13)  176 (34.11)  65 (12.60) | 32 (3.80)  188 (22.33)  48 (5.70)  7 (0.83)  3 (0.36)  140 (16.63)  15 (1.78)  2 (0.24)  25 (2.97)  5 (0.59)  14 (1.66)  5 (0.59)  2 (0.24)  61 (7.24)  3 (0.36)  35 (4.16)  7 (0.83)  24 (2.85)  31 (3.68)  63 (7.48) | 0.001  <0.001  0.648  0.622  0.117  <0.001  0.363  0.652  0.003  0.858  0.734  0.992  0.838  0.006  0.526  0.038  0.076  0.001  <0.001  <0.001 |
| **Number of comorbidities**  No comorbidities  1 comorbidity  2 comorbidities  ≥ 3 comorbidities | 403 (1.78)  437 (34.46)  278 (21.92)  150 (11.83) | 115 (22.29)  170 (32.95)  135 (26.16)  96 (18.60) | 398 (47.27)  249 (29.57)  136 (16.15)  59 (7.01) | <0.001 |
| **Sector**  Public  Private | 323 (25.47)  945 (74.53) | 98 (18.99)  418 (81.01) | 254 (30.17)  588 (69.83) | <0.001 |

p=0.108

p=0.006

Percentage of participants

|  | PWH | HIV-uninfected |
| --- | --- | --- |
| 3-months | 43 | 54 |
| 6-months | 40 | 47 |

Supplementary Figure S4: Percentage of hospitalised participants with symptoms at 3- and 6-month follow-up, by HIV status

|  | Percentage of participants |
| --- | --- |
| No symptoms | 33 |
| Up to 3 months | 17 |
| Up to 6 months | 31 |
| Onset at 3 months | 16 |
| Onset at 6 months | 5 |

Supplementary Figure S5: Evolution of symptoms among participants hospitalised with COVID-19
